# Rethinking health services around clusters of co-existing diseases: impact on integrated care for people with Multiple Long-Term Conditions

**DOI:** 10.1101/2025.05.06.25327045

**Authors:** Thomas Beaney, Jonathan Clarke, Laura Downey, Ioanna Tzoulaki, Thomas Woodcock, Mauricio Barahona, Azeem Majeed, Paul Aylin

## Abstract

**Background:** Care delivery in conventional health systems is structured around clinical anatomical specialties, often geared towards managing single conditions rather than people. As a result, people with Multiple Long-Term Conditions (MLTC) often need to see many different specialists to manage their health, resulting in fragmented care that is inefficient and costly. Certain Long-term Conditions (LTCs) frequently co-exist in identifiable clusters. Here, we examine the impact on the number of interactions with distinct healthcare services per person with MLTC if care was organised around common clusters of co-existing conditions, rather than anatomical specialties.

**Methods and findings:** We used a nationally representative sample of electronic health records from general practices in England. Patients aged ≥18 years and registered on 1^st^ January 2020 with MLTC (two or more of 212 LTCs) were included. Each LTC was assigned to one of fifteen systems representing the conventional ‘specialty-based’ model of care and to one of fifteen clusters, derived from earlier work, representing a ‘cluster-based’ model of care. We calculated the number of interactions with distinct services under each model, assuming a person has a hospital appointment for each of their conditions.

7,122,447 adults were included, with a median (interquartile range) age of 54 (39 - 67) years. Under the specialty-based model, patients would interact with a mean (standard deviation) of 3.91 (2.02) different services and under the hypothetical cluster-based model, would interact with 3.51 (1.76) different services, 10.1% lower than under the specialty-based model (p<0.001). Under the specialty-based model, 419,252 (5.9%) patients interacted with only one service, and under the cluster-based model, 594,641 (8.3%) interacted with only one service.

**Conclusions:** Hospital services organised around clusters of co-existing conditions might allow patients with MLTC to interact with fewer different services and so improve integrated care. Further work is needed to understand which specialties collaborating, and how, would have the greatest impact on enhancing person-centred care and health outcomes.

## Introduction

Modern health systems have become increasingly focused on care for individual diseases at the expense of holistic care for people. As Barbara Starfield wrote: “The benefits to health from advances in medicine in the 20th century have led to a shift away from patients’ problems to disease processes…with a demonstrated decline in focus on the person.”^1^ Conventional hospital services are organised around specialties focused on anatomical systems (such as nephrology for the renal system), rather than around combinations of conditions that often co-exist together. For people with multiple chronic conditions, commonly referred to as Multiple Long-Term Conditions (MLTC),^2^ the design of health systems around diseases rather than people, inevitably leads to an escalating number of healthcare interactions for every additional condition, particularly for those requiring hospital-based care.

There is growing evidence for the benefits of continuity of care for both patients and health systems, including relational continuity to the same clinician and continuity between different providers and specialties.^3–5^ A recent observational study in Denmark found that fragmented care, characterised by a greater number of distinct healthcare providers, or frequent transitions between providers, was associated with increased mortality and inappropriate medication prescribing.^6^ People with MLTC are at high risk of fragmented care, and so may be those who will benefit most from strategies to improve care integration.^7–9^

One of the biggest challenges to developing strategies to manage MLTC is that there is wide variation among people that have it. A study by Stokes *et al* (2021) found that in a population of 8.4 million people, there were 63,124 unique combinations of only 28 diseases,^10^ too many to effectively tailor services to each pattern. Over the last decade, much research has been conducted to identify larger groups (or ‘clusters’) of conditions that commonly co-exist within MLTC, with clusters of cardiometabolic conditions and of mental health conditions consistently found across studies.^11,12^ Examples of implementation of cardiometabolic clinics aiming to provide integrated management of both cardiovascular and metabolic conditions in ‘one-stop’ clinics suggest one route by which clusters could inform the organisation of healthcare services.^13^ However, it remains uncertain whether bringing together multi-specialty teams coordinated around the care of distinct clusters of common conditions more widely, could reduce the number of interactions with different specialist services, and so minimize the risk of fragmented care within an organization.

To address this knowledge gap, this study aims to understand how organising hospital specialist clinics which reflect clusters of conditions that commonly co-occur in people, instead of conventional anatomical-based specialties impacts on the number of interactions with distinct services per person as a measure of fragmented care. We do this applied to a large and nationally representative sample of patients registered to general practices (GPs) in England.

## Methods

### Data source and inclusion

We used the Clinical Practice Research Datalink (CPRD), a nationally representative sample of electronic health record (EHR) data from patients registered to GPs in England.^14^ We included all adult patients (≥18 years) defined by CPRD as ‘research acceptable’ (as determined by a set of quality criteria including valid dates of birth and registration^15^). We also required that patients were registered on 1st January 2020 and had MLTC, defined as two or more of 212 long-term conditions (LTCs; see full list in appendix Table A1).^2,16^ To ensure sufficient time for conditions to be coded into the healthcare record, we excluded patients who were not registered to a GP practice for at least one year before the study start date (i.e., were not registered continuously from 1^st^ January 2019 to 1^st^ January 2020).^17^ Data were linked to deciles of the 2019 Index of Multiple Deprivation, a composite index which captures domains of deprivation for small geographic units representing between approximately 1000 and 3000 people.^18^ Information on the recording of gender and ethnicity can be found in the appendix, p.2.

### Specialty and cluster assignment

Each of the 212 LTCs are assigned to one of fifteen anatomical ‘systems’, such as ‘Diseases of the Digestive System’ and ‘Skin conditions’ (Appendix Table A1). The assignment of diseases to systems was developed in an earlier study^19^ and reflects the membership of diseases within chapters in the International Classification of Diseases.^20^ These in turn reflect the medical specialties that would typically manage a condition, for example, cardiology for heart failure. We refer to this assignment as the ‘specialty-based’ model.

Our comparison ‘cluster-based’ model used the assignment of the same 212 LTCs to fifteen clusters of co-occurring diseases, derived from our earlier work (Appendix Table A2).^21^ The method of generating these clusters has been described in detail previously,^21^ but in brief, clusters were created by application of a neural-network architecture (skipgram) which learns the chronological order of conditions within each person as recorded in their EHR, to build up a quantified vector representation of each condition. We then applied a graph-based clustering algorithm to generate robust clusters, which represent conditions that tend to co-occur in one person and be diagnosed in proximity over time with other conditions in the same cluster.^21^ Earlier validation of these clusters demonstrated that they represent clinically interpretable groups which correspond to well-established associations between diseases reflected in clinical guidelines.^21^

### Statistical analysis

To compare disease assignment to clusters and systems, we used a Sankey plot to visualise the overlap of conditions. We also calculated the entropy score for each cluster to quantify how diverse the clusters are in terms of the distribution of diseases across systems. Here for each cluster, we calculate the Shannon entropy score *E*_*c*_:^22,23^

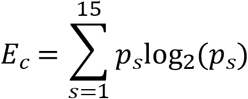

where *p*_*s*_ is the probability of each disease in a cluster being present in each system, *s*, numbered 1 to 15. A score of 0 indicates that all the conditions within a cluster are contained within only one system, and higher scores indicate more diverse assignment to different systems.

To estimate the number of interactions per person with different services under the existing specialty-based and hypothetical cluster-based models, we assumed a scenario in which each person has a hospital appointment at least once for each of their LTCs. We then calculated the number of interactions that a patient would require with different services, under each model. Given that the number of systems and clusters are identical (fifteen) this allows direct comparison and gives an upper bound on the number of distinct services a person might need to interact with, as a measure of fragmented care.

Python version 3.11.5 and Pandas version 2.0.3 were used for data manipulation and management.^24,25^ Sankey diagrams were created using Plotly version 5.16.1.^26^

### Ethics approval

Ethics approval for access to CPRD data was granted by CPRD’s Research Data Governance Process on 28th April 2022 (Protocol reference: 22_001818).

## Results

### Participant characteristics

A total of 7,122,447 adults with MLTC registered on 1^st^ January 2020 were included in the analysis (flowchart in appendix Figure A1). The median (interquartile range (IQR)) age was 54 (39 - 67) years, with 8.5% of the population aged 80 years or over (Table 1). Just over half (52.8%) of patients were female, the majority (84.1%) were of White ethnicity and there were slightly more people living in less deprived areas (51.5% in the least deprived half). The median (IQR) number of LTCs per person was seven (4 – 12).

**Table 1.**
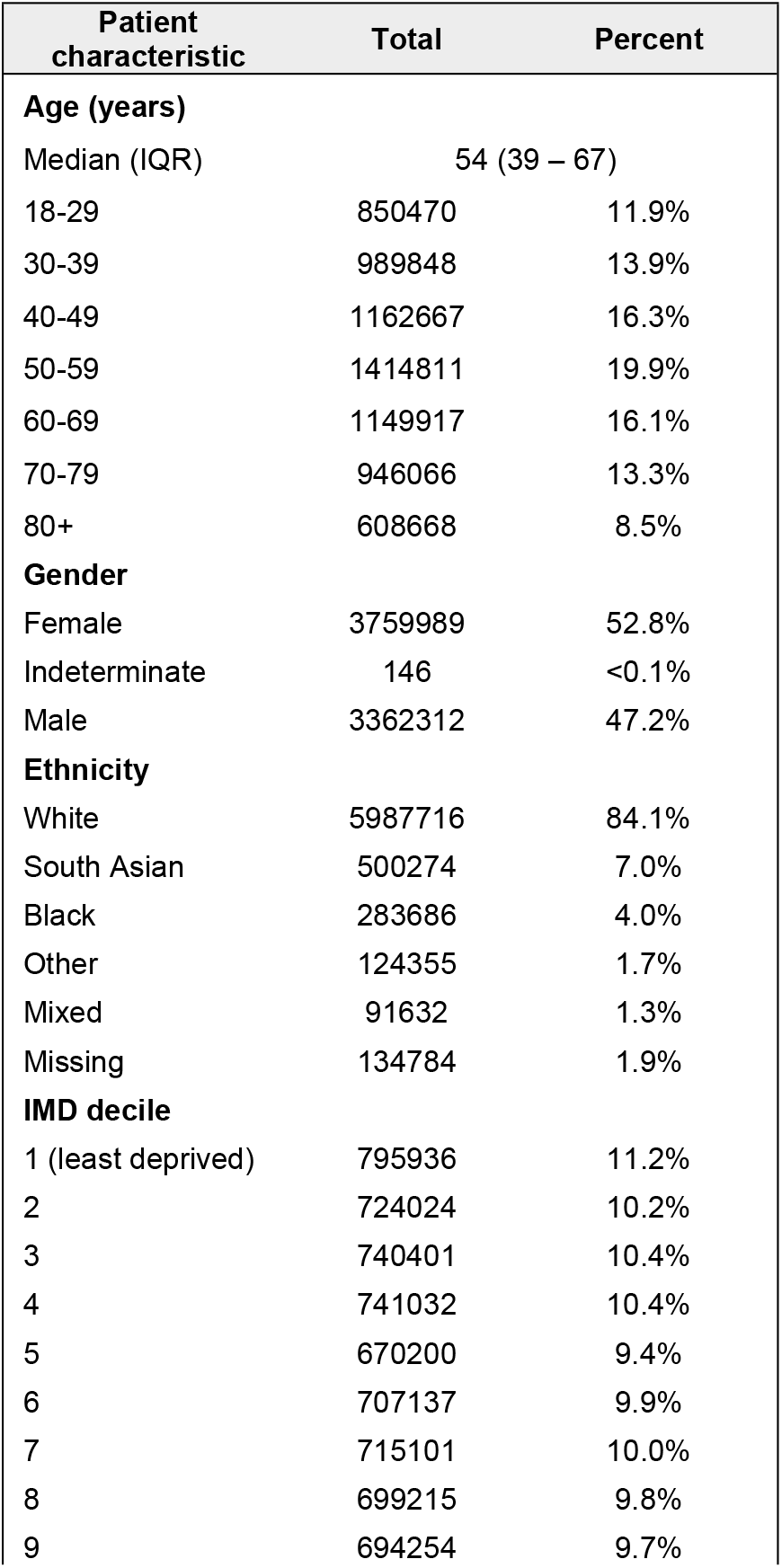
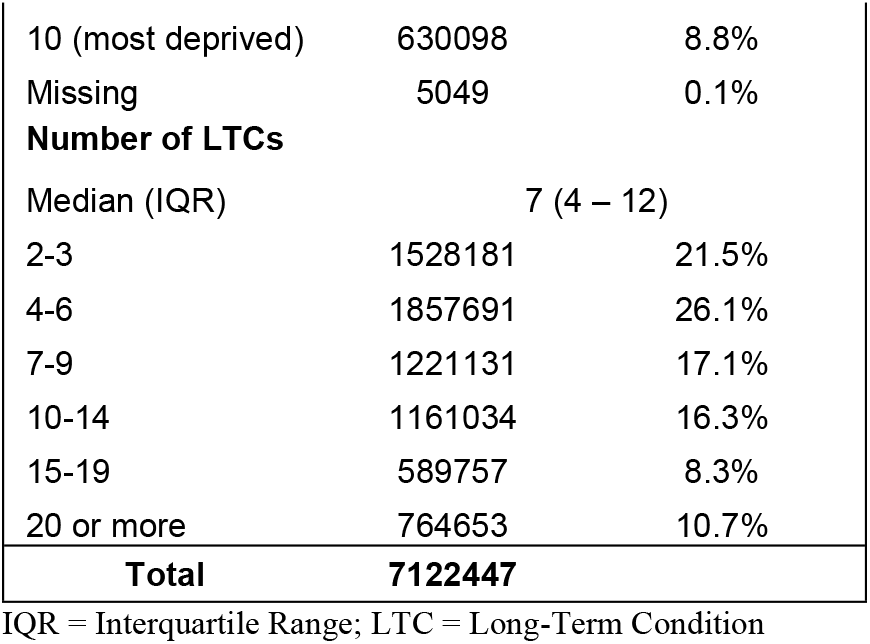
Sociodemographic characteristics of study participants.

### Assignment of diseases to cluster-based versus specialty-based models

Each of the 212 LTCs was assigned to one of fifteen systems, representing conventional hospital specialties (the ‘specialty-based’ model) and one of fifteen clusters representing conditions which co-occur (the ‘cluster-based’ model; see assignment in Appendix Tables 1 and 2). There were considerable differences in disease assignment between the two models (Figure 1). Entropy scores were calculated, giving an indication as to how mixed the clusters are with respect to the specialty-based classification. For example, the ‘malignancies’ cluster-based service exclusively contains primary and secondary malignancies, conditions which are entirely contained with the ‘cancer’ specialty service, and so giving an entropy of zero. The more heterogeneous ‘allergic, skin and pain’ cluster-based service had the highest entropy (2.49), including conditions spanning six different specialties, none of which contributed more than one third of conditions. Similarly, the ‘visual, cognitive and bone’ cluster-based service demonstrates considerable overlap of conditions managed by musculoskeletal, ophthalmology and cardiac specialties. In contrast, the ‘cardiac: arrhythmia, HF and valve’ cluster-based service had a low entropy (0.57), almost entirely contained within the ‘cardiology and cardiac surgery’ specialty service. The ‘respiratory and vascular’ cluster-based service had a higher entropy of 1.89 but with most conditions managed by a combination of respiratory and cardiovascular speciality services.

**Figure 1.**
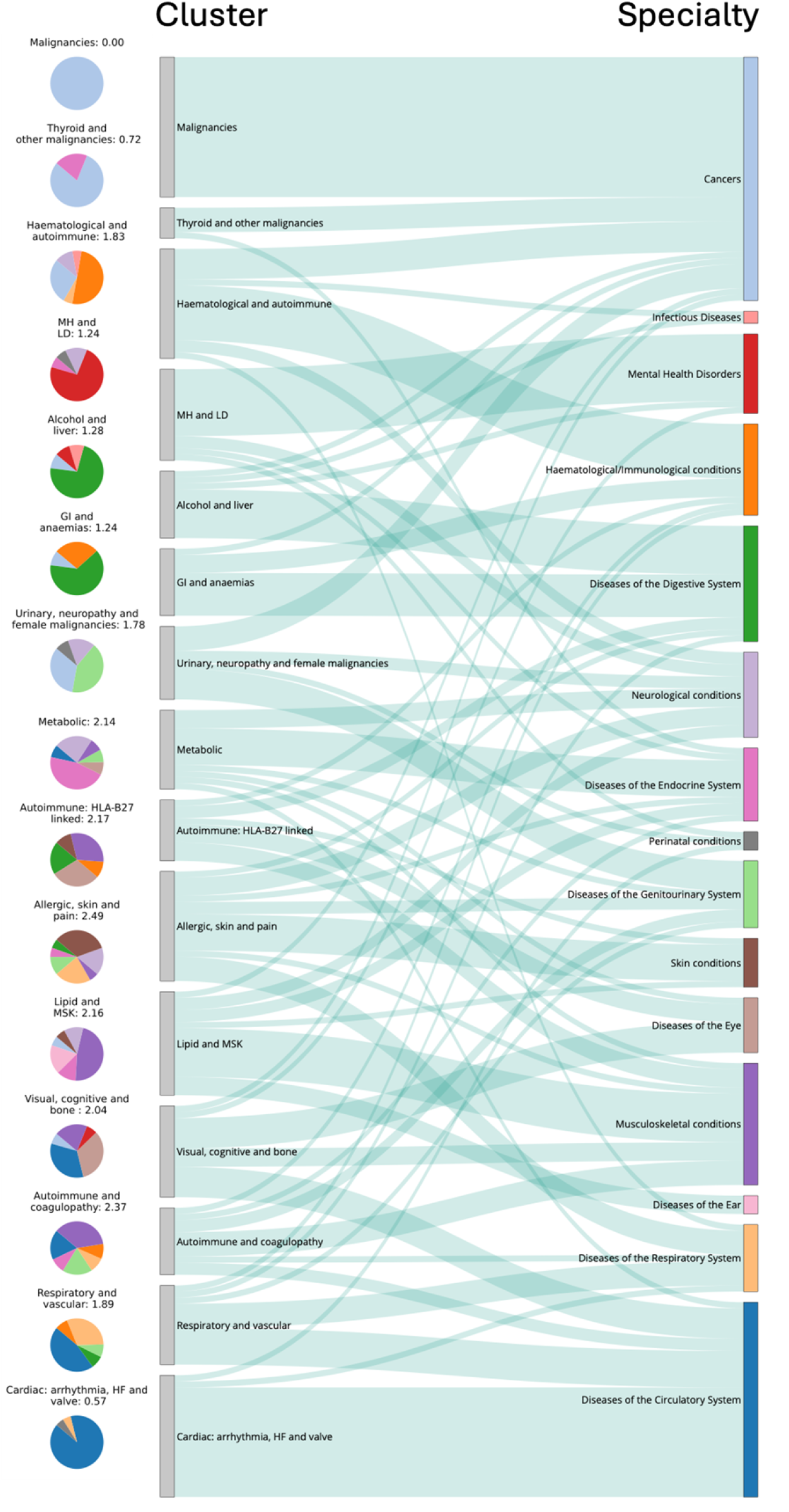
Sankey plot of the assignment of 212 LTCs to cluster-based and specialty-based models and associated entropy scores for each cluster. The grey bars on the left represent clusters and the coloured bars represent specialties; the connections between them reflect the number of diseases that are shared between the cluster and specialty. The pie charts on the left represent the percentage of diseases within each cluster that are present in the specialties, coloured according to the specialties, with the associated entropy value for each cluster. The assignment of individual diseases within each cluster and system are given in Appendix Tables 1 and 2.

### Interactions with different services per person

Assuming each patient had one hospital appointment for each of their LTCs, under the existing specialty-based model, patients would interact with a mean (standard deviation (SD)) of 3.91 (2.02) different services per year (Table 2). Under the hypothetical cluster-based model, where conditions are assigned to one of fifteen clusters, patients would interact with a mean (SD) of 3.51 (1.76) services per year, 10.1% lower than under the specialty-based model (p<0.001).

**Table 2.** Number of interactions per person with distinct specialty-based versus cluster-based services.

|  | Specialty-based (N=15) |  | Cluster-based (N=15) |  |
| --- | --- | --- | --- | --- |
| **Mean** | 3.91 |  | 3.51 |  |
| **Standard deviation** | 2.02 |  | 1.76 |  |
| **Median** | 3 |  | 3 |  |
| **Lower quartile** | 2 |  | 2 |  |
| **Upper quartile** | 5 |  | 5 |  |
| **Total interactions** | **Number** | **Percentage** | **Number** | **Percentage** |
| **1** | 419252 | 5.9% | 594641 | 8.3% |
| **2** | 1673041 | 23.5% | 1833847 | 25.7% |
| **3** | 1477165 | 20.7% | 1618387 | 22.7% |
| **4** | 1188509 | 16.7% | 1244196 | 17.5% |
| **5** | 891107 | 12.5% | 844906 | 11.9% |
| **6** | 626625 | 8.8% | 510496 | 7.2% |
| **7** | 408924 | 5.7% | 274521 | 3.9% |
| **8** | 239014 | 3.4% | 129055 | 1.8% |
| **9** | 123120 | 1.7% | 51380 | 0.7% |
| **10 or more** | 75690 | 1.1% | 21018 | 0.3% |

Under the specialty-based model, 419,252 (5.9%) patients interacted with only one service and 5,030,154 (70.6%) interacted with three or more services. Under the cluster-based model, 594,641 (8.3%) interacted with only one service, and 4,693959 (65.9%) interacted with three or more services (Table 2). Of all patients, 2,945,127 (41.4%) had fewer interactions with different services under the cluster-based model than the specialty-based model, 2,973, 911 (41.8%) had the same number of interactions and 1,203,409 (16.9%) had more interactions.

### Interactions with different services according to long-term conditions

There was large variation in the number of interactions with different services per person according to underlying conditions (Appendix Table 3). Under both models, people with hyperkinetic disorders had the fewest interactions with unique services. Under the specialty-based model, people with diabetic neuropathy had the most interactions, and under the cluster-based model, those with pulmonary fibrosis had the most. For 198 (93.4% of the 212) conditions, people with each condition on average interacted with fewer services under the cluster-based model than the specialty-based model. The largest reductions in unique service interactions under the cluster-based model were seen for people with diabetic neuropathy, Parkinson’s disease and Meniere’s disease. Conditions associated with older age, including dementia, spondylosis, cataracts and osteoarthritis also had large reductions in unique service interactions per person. For only fourteen (6.6%) conditions, people with the condition interacted with more unique services under the cluster-based model. This included people with thyroid disease, alcohol misuse, rheumatoid arthritis and schizophrenia.

## Discussion

A common challenge for people with MLTC is the frequent need to see different specialists to manage their conditions, resulting in fragmented care.^27^ This is linked to difficulties navigating healthcare, lower patient satisfaction and poorer health outcomes.^6,28^ Our findings indicate the potential improvements to integrated care that could be achieved for people with MLTC if services were organised around conditions that commonly co-exist in people, rather than conventional anatomical specialties. Under a cluster-based model of care, patients interacted with 10% fewer distinct services on average, with 41% of patients interacting with fewer and only 17% interacting with more services. Notably, people with conditions associated with older age, including dementia and osteoarthritis, or cardiometabolic conditions, such as diabetes and its complications, demonstrated larger reductions in use of multiple services. Reducing the number of different services a person interacts with would minimise the risk of fragmented care and could improve integration across hospital outpatient services, thereby promoting person-centred care.

Although our analysis represents an idealised scenario in which a patient has a specialist appointment for each condition, it indicates the potential impact of reorganising health services to reflect how conditions co-exist within people. Beyond the impact on reducing fragmented care, it could have wider benefits, supporting earlier diagnosis and collective management of conditions in the same cluster. This could improve efficiency and lead to cost savings, allowing specialist services to be re-allocated elsewhere; for example, to reduce wating times for specialist assessment.

### Existing evidence supporting clusters in healthcare services

Despite many studies generating clusters of diseases or people in the context of MLTC, there are few examples of how insights from clusters can be used to improve patient care. There are several routes by which disease clusters could inform models of care delivery, without requiring fundamental redesign of services, including co-locating clinics and forming multi-specialty clinics. Community or hospital services could co-locate and synchronise specialist clinics guided by the mix of specialties usually managing conditions within clusters, thereby reducing the need for patients to attend separate clinics on different days. For example, given the large reductions in number of interactions under a cluster-based model for conditions associated with older age, co-locating dementia, ophthalmology and musculoskeletal services might reduce the burden of repeated hospital visits for people with these conditions which commonly co-occur in the ‘visual, cognitive and bone’ cluster.

Evidence from co-locating health and social care services suggests it can promote service innovation and effectiveness,^29^ while co-locating mental health services in community spaces was found to improve access and reduce stress related to transitions between services in the same space.^30^ Within primary care, although co-location can increase formal and informal discussions between staff, alone it may not be sufficient to ensure collaboration and integration of care, also requiring continuity of staff and shared understanding of different roles.^31^

Alternatively, specialties could collaborate in multi-specialty teams, which collectively manage the conditions within a cluster. The British Society of Gastroenterology, for example, have advocated for the creation of ‘alcohol care teams’, combining hepatology or gastroenterology services alongside addiction and psychiatric services,^32^ aligning with the co-membership of conditions managed by these services within the ‘alcohol and liver’ cluster in our work. Cardiovascular and metabolic conditions are frequently found to cluster together across populations,^11^ and there are several examples of the implementation of cardiometabolic clinics seeking to manage these conditions concurrently.^13^ The rationale for these joint clinics is supported by our results, with conditions in the ‘metabolic’ cluster, such as diabetes, diabetic complications and chronic kidney disease demonstrating large reductions in interactions with different services under the cluster-based model. However, there is currently a lack of robust evidence for the effectiveness of multi-specialty clinics for cardiometabolic conditions and wide variation in how such clinics are implemented.^13^

### Implications for health services

The insights offered by clusters could help to design interventions and prioritise resources for those clusters which are at greatest risk of fragmented care. For example, the ‘allergic, skin and pain’ cluster was found to include conditions managed by many different specialties (Figure 1) and so might benefit from strategies to improve collaboration across these specialties. Co-location and multi-specialty teams are promising approaches for addressing the challenges of MLTC, but their implementation faces challenges including workforce availability and collaboration between specialties and professions. Engaging stakeholders early in the planning process, including patients, healthcare staff and policymakers is essential to ensuring that changes are clinically aligned and address patient needs, particularly when guided by data-driven approaches such as clustering.

Rigorous evaluation of any intervention is also critical and should be built into the design from the outset. This includes understanding both short-term outcomes (e.g., blood pressure) and long-term outcomes (e.g., mortality), as well as measures of patient satisfaction, healthcare worker satisfaction, cost-effectiveness and equity impact. This would be aided by future research establishing the metrics of quality of care that matter most to people with MLTCs.

The potential success of a cluster-based model hinges on its integration with primary care where GPs aim to provide person-centered continuity of care.^33^ Recent evidence suggests that there is no association between continuity in primary and secondary care for people with MLTC,^34^ highlighting the need to develop strategies to improve continuity across primary and secondary care. Bridging cluster-based secondary care with GP-led coordination could enhance care integration by aligning specialized hospital services with the holistic oversight provided in general practice. Used in this way, and by collaborating better with primary care services, insights from clusters can support health systems to move away from rigid single-disease pathways to models of care that more closely align with the complex needs of people with MLTC. Supporting this requires expansion of the primary care workforce, including GPs, while use of additional staff such as care navigators could also act as potential solutions to improve integration across services.^34^

### Strengths and limitations

A strength of this study is its application to a large dataset which is representative of the English population, enhancing its generalisability.^14^ We used the assignment of a broad set of 212 LTCs to fifteen clusters which were identified from our earlier work, and found to correspond to clinically meaningful groupings of conditions.^21^ However, the ‘system’ categorisation assumed for the existing specialty-based model is an imperfect match for specialties. For example, ‘Diseases of the Circulatory System’ includes conditions that may be managed predominantly by vascular surgery services (e.g., peripheral vascular disease) or by neurology or stroke services (e.g., stroke subtypes). While alternative categorisations could be used, such as the referral specialty there are many more referral specialties than clusters, which would have required a method of combining or merging specialties to create a comparable number of referral specialties to clusters. A further challenge is that there is local and regional variation in the specialty to which people may be referred for a given condition, for example, spinal problems may be referred to either orthopaedics or neurosurgery. Hence, our findings are indicative, rather than giving an absolute measure of the services a person with MLTC would interact with.

Clustering is not the only approach by which to identify how service integration could be optimised. For example, identifying groups of services which are commonly used overall in the population, such as ophthalmology and orthopaedic services (which individually account for many total hospital outpatient attendances) might help target services for co-location and reducing healthcare burden for patients, independent of conditions managed by these services clustering together. Future research could investigate which approaches for identifying service collaborations would have the greatest impact on reducing the need for multiple visits and fragmented care.

A further limitation is our assumption that patients have an appointment for each of their conditions, given that severity and impact of conditions is likely to vary over time, affecting the chance of needing specialist input. Applying a weighting according to the likelihood of requiring a hospital appointment for a condition could be used to down-weight conditions for which patients are more likely to self-care. However, within our data it is not possible to robustly identify the condition prompting a referral. Furthermore, any weighting would complicate interpretation, as if services were organised differently, there would inevitably be changes to how patients are referred and receive ongoing care. This again highlights the need for ongoing evaluations to monitor the impact of any changes to models of care.

## Conclusion

We found that organising hospital services around clusters of conditions that commonly co-exist in people with MLTC, rather than around conventional anatomical specialties led to a 10% reduction on average in the number of distinct services a person needs to see, which might reduce fragmented care and improve service efficiency. Given broader health policy goals of achieving more integrated care, clusters may act as a guide for rethinking how we deliver care, away from diseases and towards more person-centred health services. Further work is needed to understand which specialties collaborating, and how, would have the greatest impact on improving integrated care. Successful implementation of a new model of care would also require addressing challenges such as how best to co-locate services and form multi-specialty teams.

## Data Availability

Data from CPRD are not publicly available to to protect participant anonymity. Data are available to researchers meeting certain requirements, as described here: https://www.cprd.com/research-applications

## Acknowledgements

Data management was provided by the Big Data and Analytical Unit (BDAU) at the Institute of Global Health Innovation (IGHI), Imperial College London. Thanks to Mark Cunningham for assistance with extracting data from CPRD.

